# The World Is Completely Unprepared For Detecting Emerging Viruses/Bacteria In The Air

**DOI:** 10.1101/2022.08.09.22278555

**Authors:** Devabhaktuni Srikrishna

**Affiliations:** Patient Knowhow, Inc.

## Abstract

There is an immediate need for efficient networks to detect and control novel/emerging bioaerosol threats. In 2022, the Pentagon Force Protection Agency (PFPA) found detection of emerging bioaerosol threats to be “not feasible for daily operations” due to cost of reagents used for metagenomics, cost of sequencing instruments, and time/cost for labor (subject matter expertise) to analyze bioinformatics. If the Pentagon experiences these operational difficulties, they may also extend to many of the 280,000 buildings (2.3 billion square feet) at 5000 secure US DoD military sites, 250 US Navy ships, and beyond in civilian buildings around the world. These economic barriers can still be addressed in a threat-agnostic manner by dynamically pooling samples from a network of dry filter units (DFUs) to detect spatio-temporally correlated “spikes” in novel pathogen concentrations, termed Spike Triggered Virtualization (STV). In STV, pooling and sequencing depth are automatically modulated in the next cycle in response to novel potential biothreats in the sequencing output of the previous sequencing cycle. By running at a high pooling factor and lower depth unless triggered by a potential pathogen spike, average daily and annual cost per DFU can be reduced by one to two orders of magnitude depending on chosen trigger thresholds. Artificial intelligence (AI) can further enhance sensitivity of STV triggers. Risk of infection remains during the 12-24 hour window between a bioaerosol incident and its detection, but can in some cases be reduced by 80% or potentially more with high-speed indoor air cleaning exceeding 10 air changes per hour (10 ACH) similar to passenger airplanes (Airbus A319, A321 and a Boeing 737-Max8/9) that were tested in flight. Costs of 4 ACH or higher are expected to rise non-linearly with ACH using central HVAC systems and be cost-prohibitive. Whereas 10 ACH or more can be achieved economically by recycling the air in rooms with low-noise, portable air filtration systems tested herein for which costs scale linearly with ACH. For typical ceiling heights (< 10 ‘), the cost per square foot cost for low-noise air filtration exceeding 10 ACH was found to be approximately $0.5 to $1 for Do-It-Yourself (DIY) and $2 to $5 for HEPA.

## Introduction

The Pentagon Force Protection Agency (PFPA) has been at the forefront of innovations in threat-agnostic, bioaerosol security operations [72] such as use of barcoded DNA for mapping indoor aerosol transport between dry filter units (DFUs) in 2016 [1] [2]. On March 2, 2023 the PFPA together with the Countering Weapons of Mass Destruction Office (CWMD) of the Department of Homeland Security (DHS) jointly issued a request for information (RFI) on research and development for “Rapid Assay Biosensor Redesign” [74]. The main purpose of the RFI appears to be to improve public safety by detecting bioaerosol threats in real-time, and it mentioned not only traditional biological weapons agents (BSAT) but also emerging and unknown (novel) biological threats. The desired requirements included rapid detection of fixed targets at high-accuracy (< 5 minutes, BSAT with expandable target assays, PCR-like sensitivity, near-zero false positives). Even if every desired requirement were to be met there remain 3 countermeasures which adversaries could anticipate. First, fixed targets fail with unknown pathogens lacking validated targets, as noted by GAO (page 40 of [75]). Significant advance notice is typically needed for library expansion and then validation of new targets. In recent years many biothreats were novel and contagious, not on the shortlist of BSAT (e.g.

SARS-CoV-2). For instance, SARS-CoV-2 spread for weeks undetected (cryptically) through Washington state during early days of the pandemic before initial detection in hospitals was later documented [76] [77]. Such undetected contagion could easily happen with a new virus e.g. at the Pentagon. Second, even if detection is within 5 mins of release of bioaerosols, people can be infected in that time. Human reaction time to “turn on” protection in response to detection events (e.g. use respirators, air filtration, or UV) can be longer than 5 minutes. Third, even if pathogens (BSAT or NBT) are detected, the “treat” in detect-to-treat can be unreliable, unavailable, delayed, or unsafe (as discussed in further detail below). Like roofs on a rainy day, 24×7 protection from pathogen exposure (no delay) is needed to ensure safety whether 5 mins, 60 mins, or 24 hours.

On June 28, 2022, PFPA presented its experience with detection of emerging/novel bioaerosol threats (NBT) as being “not feasible for daily operations” [3]. These can include the universe of artificial-derived (synthetic or engineered) pathogens or natural pathogens for example those with pandemic potential such as most recently SARS-CoV-2 or H5N1 [70] in the future. If the Pentagon experiences such operational difficulties, they may also extend to many of the 280,000 buildings (2.3 billion square feet) at 5000 secure DoD military sites, 250 active Navy ships at sea [5], and beyond in civilian buildings around the world. PFPA currently conducts biothreat testing for mail and environmental samples 24/7/365 at an onsite BSL 2/3 lab. PFPA surveils routinely from aerosol samples collected from a network infrastructure of DFUs located across the Pentagon reservation to track aerosolized biothreats such as one that might be released from an improvised bioaerosol device (e.g. a perfume bottle) or through aerosol contagion among people inside the Pentagon. PFPA is using molecular PCR assays and immunoassays to detect the presence of Biological Select Agents and Toxin (BSAT) known in advance, which are sensitive, specific, timely and within budget. However the Pentagon lacks a similar capability for NBT due to the tough economics of gene sequencing technologies necessary for real-time, accurate detection of aerosolized pathogen species, variants, and single nucleotide polymorphisms (SNPs) needed to differentiate viruses/bacteria between human-threatening (true positive) versus animal or plant (false positive).

For public safety against all bioaerosol threats, a robust, future-proof solution is needed, complementary to target-specific biodetection. Below we discuss a path to make metagenomic sequencing feasible for daily detection of novel bioaerosols using virtualization and artificial intelligence (AI), and we identify cost-effective, 24×7×365 air cleaning options to bridge the gap between the bioaerosol release and its initial detection up to 24 hours later.

### Pentagon Pilot Study of Metagenomics (2022)

From March 2021 to March 2022 the Pentagon piloted metagenomic sequencing technologies to characterize the environmental background, and additionally to evaluate it as a potential threat-agnostic method to detect emerging and advanced biological threats [3]. A total of N=105 DFUs located indoors and outdoors were sampled once a month. The metagenomics revealed the “microbial background community is large and complex: it contains many different species of naturally-occurring and human-associated bacteria and viruses.” In some instances, samples contained high levels of fungi and human DNA which complicated analysis. In conclusion, PFPA found the metagenomic method was “not feasible for daily operations” due to its excessive cost, time consumed, large quantities of genetic data requiring high computational capability, and subject matter expertise (SME) to analyze the data. Furthermore, the Pentagon highlighted several other operational challenges including bioinformatic biases, microbial background in sampling devices, and high abundance organisms inhibiting detection of low abundance organisms. The Pentagon found bioaerosol metagenomics of the environment can be useful to improve its PCR assay design, but further technological advances are needed to reduce both the cost of resources and ambiguity of results in order to make metagenomics feasible as a threat-agnostic method to identify emerging threats. When I asked what are the costliest and most time consuming elements of the solution, the Pentagon specifically identified the reagents used for metagenomics are two orders of magnitude more costly than PCR, in addition to the cost of sequencing instruments (machines) and the labor (expertise) to analyze the bioinformatics.

For environmental background sampling applications seeking to examine the microbes captured in each DFU, emerging pathogen detection turned out to be cost-prohibitive in the Pentagon pilot. Although the density of novel pathogens in the air may be fewer than 1000 per cubic meter [6] it may also contain 10 million other microbes or more [7]. As many as 10,000 microorganisms may need to be sequenced to find one novel human pathogen. A DFU collecting air typically at the rate of 1 cubic meter per minute can easily result in several hundreds of millions of microbes in each sample. Depending on the desired breadth and depth of the sequencing (quality), the cost of a single run of short-read sequencing can easily exceed $1000 [8] generating hundreds of gigabytes of data to be analyzed [9].

It is important to note that during the Pentagon pilot [1] the DFU samples were collected and analyzed monthly and there were only N=105 DFUs which is small considering the size of the Pentagon (6M sq. ft.). As such, the PFPA pilot was not able to fully test metagenomics for detecting novel pathogens in air by observing daily/weekly trends (1st derivative) of bioaerosol concentration in pooled DFU samples, e.g. such spikes can also be seen weekly for COVID in aggregated wastewater data in as shown in Figure 1 [79].

**Figure 1:**
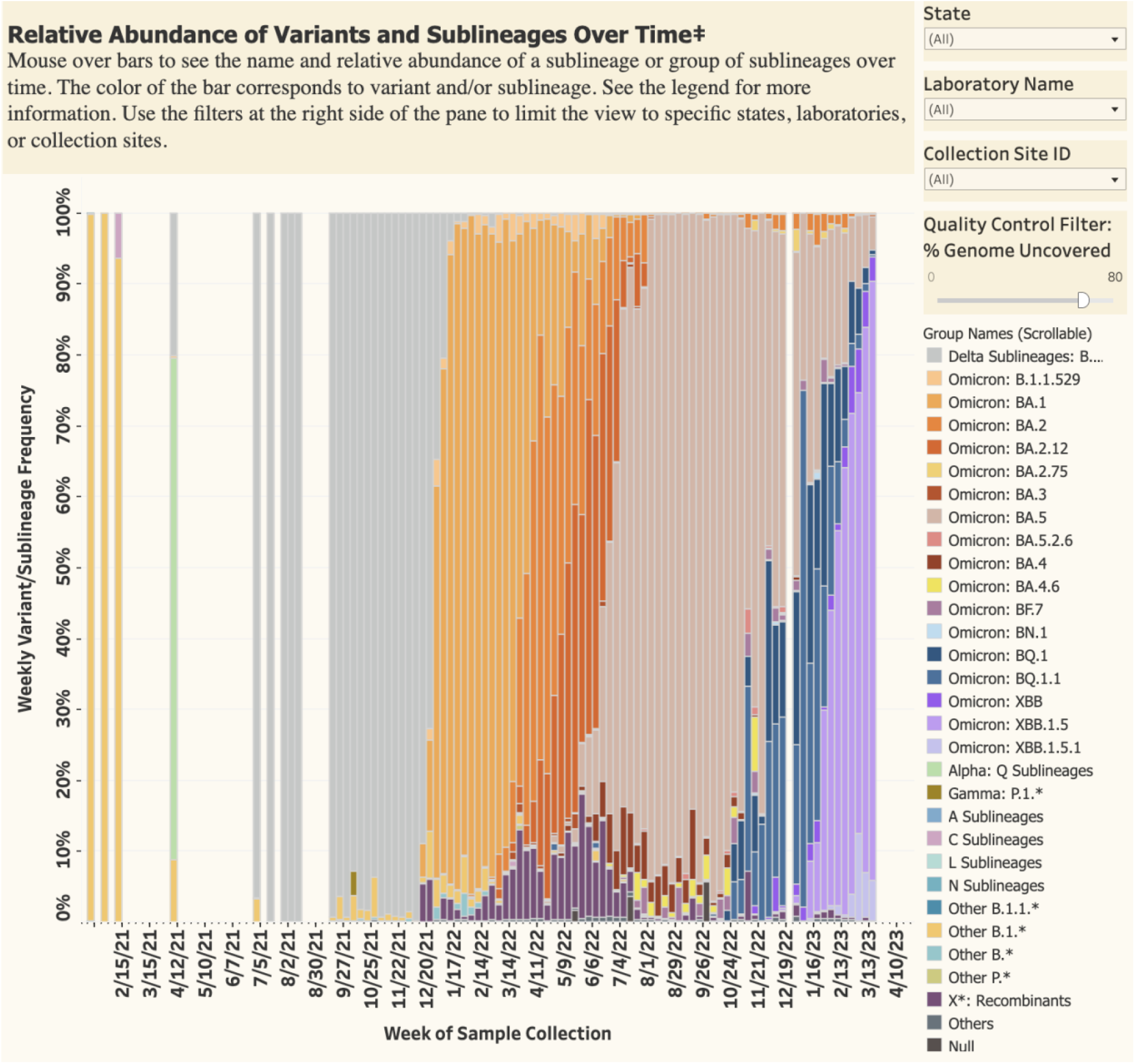
Relative Abundance of Variants and Sublineages Over Time (3/28/2023, FDA)

During the pandemic, wastewater has typically been aggregated and run through metagenomics on a weekly/daily basis. In principle, something similar can be done to improve economics of air sampling by pooling multiple DFUs together on a daily basis (“waste-air”) across the Pentagon campus to observe the trend (1st derivative). Rapid rises if seen for a specific species or variant would be an early warning if there ever was to be a virus or bacteria in the air growing unexpectedly. Upon seeing such a warning signal, an increased rate/depth of sequencing would then be needed to confirm (e.g. SNP) and drill down the location to find out which DFU on the Pentagon campus it is coming from (typically not easy to do with wastewater) as discussed in the next sections.

### Spatio-temporally correlated “spikes”

Detection of the smallest quantities of a pathogen may be needed for environmental background sampling. If that stringent requirement is relaxed, aggregation or “pooling” of a larger number of DFUs in the network across fewer sequencing machines can slash the average daily cost and time within a budget targeted (by PFPA). As explained in the Air Force tactics for bioaerosol collection and identification [10], the operational benefits of pooling a large number of DFUs to be analyzed at once include a substantial reduction in expenditures on consumable materials, increased sample throughput, and lower total analysis times. However, pooling also theoretically increases the risk of a false negative due to sample dilution. If gene sequencing all samples to retain full sensitivity is cost-prohibitive, how much pooling can still reduce risk by enabling daily operations without excessive loss of sensitivity?

A “spike” in concentrations across the DFU network can be a cost-effective signal for a potentially novel pathogen. The Air Force tactical doctrine [10] also points out that although it is unlikely, a covert aerosol release of a biological agent would only be captured by a single DFU, the possibility exists. The risk of this possibility can be mitigated with deployment of more DFUs in the network. Since it is too expensive to use metagenomics to analyze all DFUs every day to search for pathogen signatures (e.g. BSAT) at PCR-like sensitivity, the alternative is to detect unexpected “spikes” or sharp rise in concentrations across multiple DFUs (locations). Although mixing or pooling ‘N ‘ DFUs can result in slightly lower sensitivity, this can be offset by detection of “spikes” in bioaerosols (> N times higher concentration than original minimum sensitivity level). This can be a cost-effective tradeoff if the pooled DFU network density is high enough to capture emissions near the source (or point) of bioaerosol release. In case of contagious pathogens exhaled by infected people such as SARS-CoV-2, spatially localized “spikes” of bioaerosol can be observed not only as a result of a rising number of infected people (“waves”) but are likely because bioaerosol emissions can vary by one or two orders of magnitude due to physiological differences among infected people [6] [11]. For example, temporally correlated spikes across an enterprise (UCSF hospital, UCSF-ATPR) were measured by diagnostic test positivity among people infected asymptomatically with SARS-CoV-2 in Figure 3 of [80]. Spikes closely tracked rise and fall of test positivity within its city over two years (San Francisco, SF-ALL) and ancestral, Delta, Omicron waves can be clearly seen.

### Spike Triggered Virtualization (STV)

Unlike PCR, the gene sequencing process introduces errors into the reads of a pathogen ‘s sequence but the error rate can be overcome by increasing the “depth”: the number of times a given nucleotide in the pathogen ‘s genome has been sequenced (from repeated instances) in the sample. At a lower depth it may be possible to confidently recognize the species of an emerging pathogen but not necessarily individual mutations within that species. At higher depth the confidence in the characterization of the novel pathogen can be increased by more accurately recognizing features of that pathogen ‘s genome including as single-nucleotide polymorphisms (SNPs) which have been implicated in increased pathogenicity or transmissibility of viruses [12] [13] [14]. Once a “spike” at a chosen threshold concentration in the air indicates the presence of a potentially novel bioaerosol, to increase confidence it becomes necessary to confirm the presence of the novel pathogen at higher depth and potentially also narrow its specific filter location. This can in turn be done by reducing the pooling factor in subsequent sequencing of DFU samples, increasing the cost per sequencing run in response to the detection of the spike.

Sharing a computer to concurrently run multiple tasks (in this case also gene sequencing machines) and dynamically adjusting the resources up or down as needed by a task is known as “virtualization” in computer science. After a “spike” in RNA/DNA is initially discovered in a pooled sample at a low depth chosen to control daily costs, it is necessary to sequence at greater depth and reduce pooling in subsequent cycles termed “Spike Triggered Virtualization” (STV). The higher rate of sequencing is to rule out false positives, increase confidence in adopting temporary precautions (e.g, N95, faster filtration, UV etc), and narrow down specific DFU locations. STV is illustrated in the model (Figure 2) showing how the maximum allowable pooling factor (per sequencing run or per machine), sequencing cost per filter, and annualized cost for the Pentagon pilot study would vary as a function of minimum pathogen concentration and minimum sequencing depth of the trigger. By running at a high enough average pooling factor in the absence of spikes, the daily and annual cost per DFU can be reduced by one to two orders of magnitude (Figure 2) depending on the trigger thresholds chosen.

**Figure 2:**
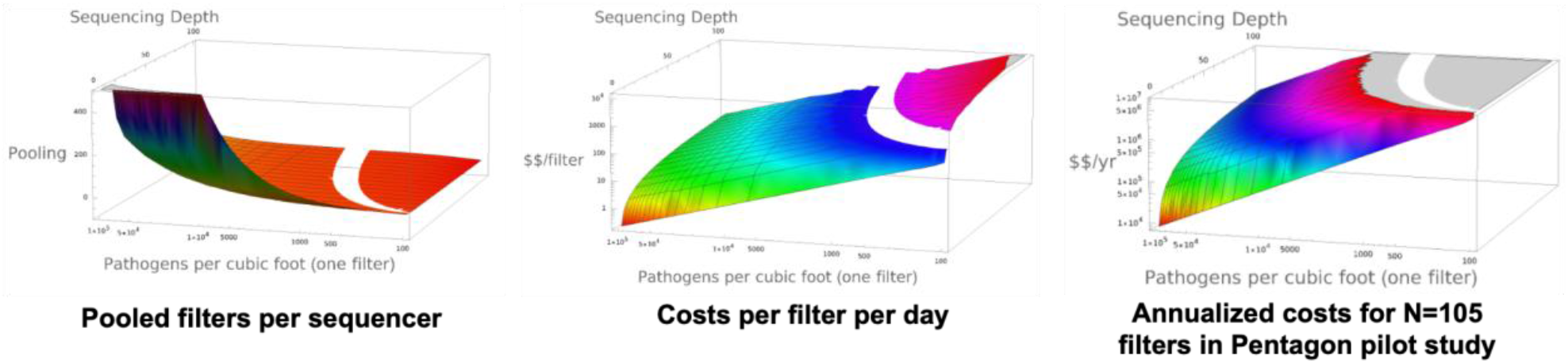
Spike Triggered Virtualization (STV)

The pooling factor and costs are computed based on certain model assumptions (Table 1) with pathogen density and depth varying (Figure 2). Although model assumptions (Table 1) can vary, the shape of the curves (Figure 2) is similar.

**Table 1:** Model for pooling multiple filters.

| Quantity | Symbol | Formula | Value | Units |
| --- | --- | --- | --- | --- |
| Genetic material density | V |  | 1,000,000 | per cubic foot |
| Air Flow rate | A |  | 300.00 | cubic feet per minute |
| Genetic material flow rate | F | $V * A$ | 300,000,000 | per minute |
| Average genetic material length | L |  | 20,000 | bases |
| bases flow rate | Fb | $F * L$ | 6,000 | Gigabases per minute |
| Bases per sequenced per run | B |  | 1 | Gigabases |
| Genetic material sequenced per run | S | $B / L$ | 50,000 | Genetic material |
| Genetic material sequencing cost per run | Cr | | 1,000 | \$ per run |
| Target pathogen density | $V_{t1000}$ | | 1,000 | per cubic foot |
| Total genetic material to pathogen ratio | H |  | 1000 |  |
| Minimum detected # of copies to identify it (sequencing depth or coverage) | $Id_{10}$ | | 10 | |
| Bases sequenced to identify it | $Bid_{10}$ | $V * Id_{10} * L / V_{t1000}$ | 200 | Megabases |
| Maximum permissible pooling factor to identify it | $P_{10}$ | $B / Bid_{10}$ | 5 | |

As the desired depth increases further, pooling may not be feasible at all pathogen concentrations and instead, multiple sequencing runs (or machines) become necessary per each filter. This is depicted as a negative pooling factor in the first chart (Figure 2). As with Table 1, although model assumptions (Table 2) may vary the shape of curves (Figure 2) remains similar.

**Table 2:**
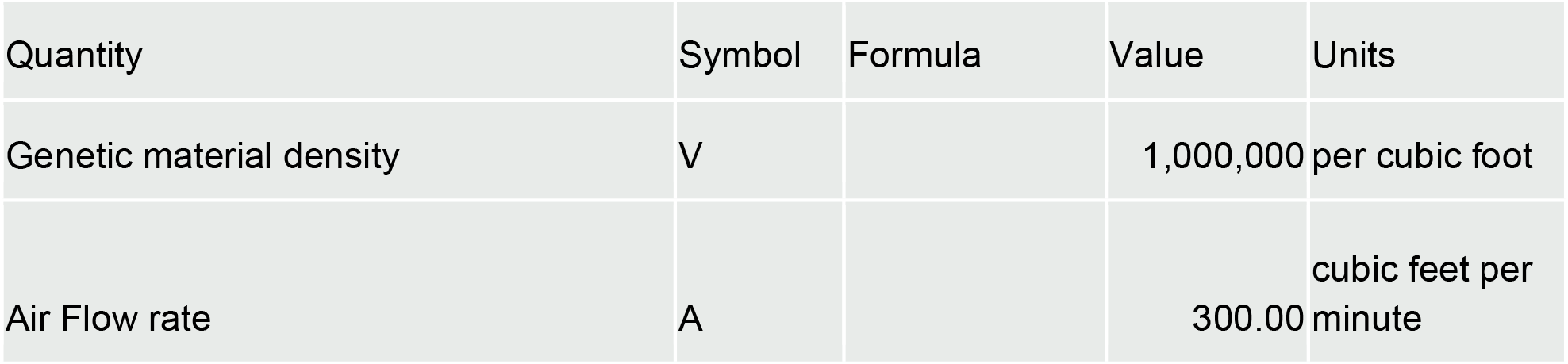
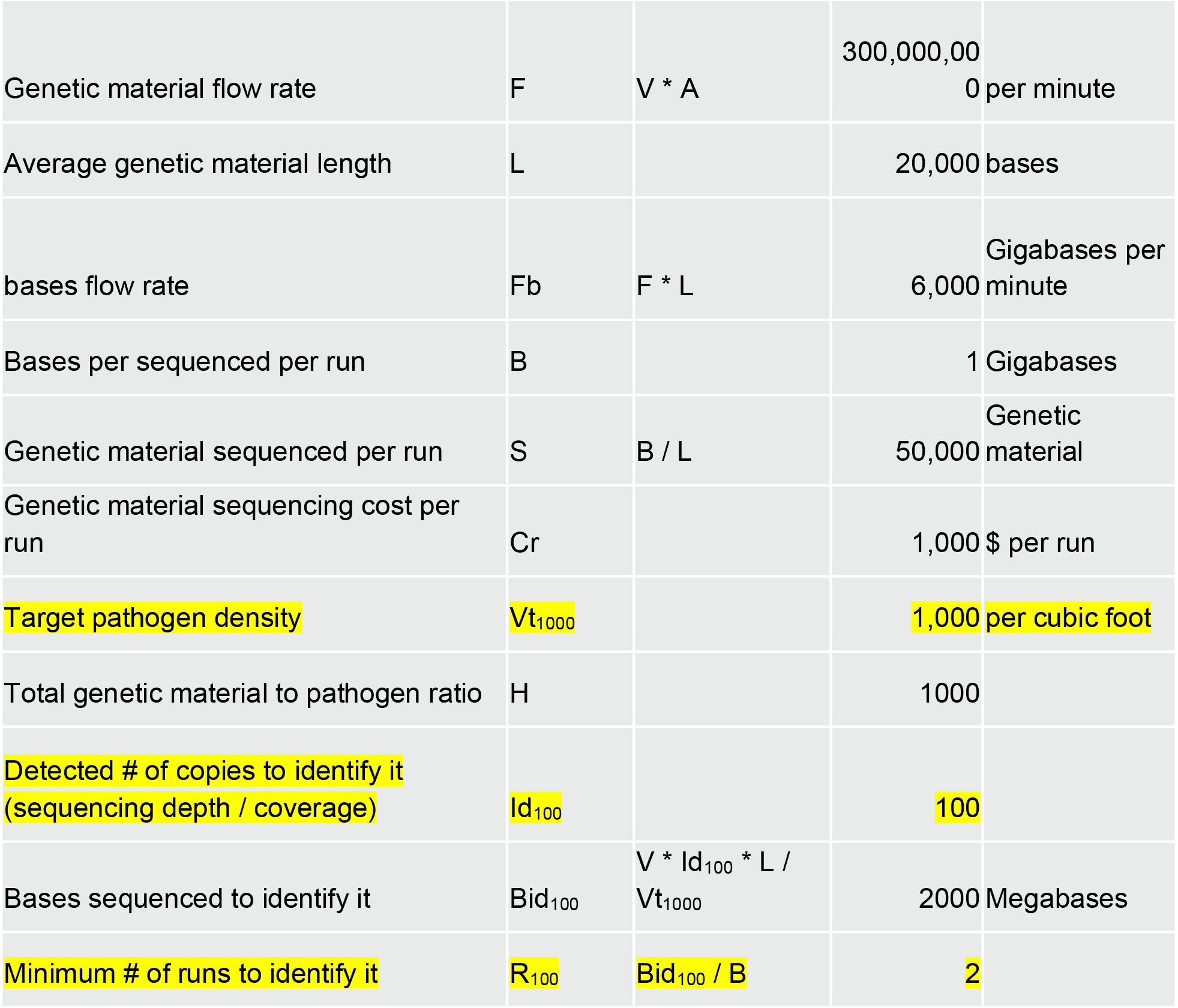
Model for number of sequencing runs per filte.

### Artificial Intelligence (AI)

AI can further enhance sensitivity of STV triggers. The sensitivity of the trigger for detection of pathogenic novelty in STV depends on the chosen threshold concentration to detect the spike. Higher concentration relative to expected levels increase confidence in the detection of novelty (e.g. “reference normalized reads” used by the Pentagon). However to maintain efficiency and reduce daily cost these would need to be further filtered to exclude viruses that affect other kingdoms (e.g. plants) which in some studies comprised the bulk of sequenced microbes [7].

There are a few opportunities to enhance the sensitivity of the STV trigger with artificial intelligence (AI). First, some suggest AI can be trained to recognize mutants of cross-kingdom viruses that are more likely to affect humans such as influenza [17]. In the pilot study, the Pentagon ‘s contractors used pathogen databases such as BLAST, PanGIA, and Kraken to recognize novel pathogens and tell them apart from other pathogens that primarily affect plants and animals. However novel human pathogens may not be entirely captured in these databases and in some cases can be cross-kingdom (e.g. plant viruses that can infect humans) [18]. For example the AI may be trained to recognize sequences that potentially match human cell entry receptors [19]. Second, AI can be trained to more robustly recognize novel pathogens present with artifacts from the sequencing process such as multiple genome segments incompletely assembled with sequencing errors.

### Subclinical Person-to-Person Contagion (P2P)

PFPA recommends masks for high community levels [20] in spite of available measures such as ventilation, filtration, and ultraviolet (UV) light disinfection [73] for long-range risk reduction.

Even with good ventilation, people socializing at close proximity (e.g. at lunch < 3 feet for 30 minutes) or who have fleeting contact in hallways are at risk for pathogen transmission [21]. Multiple case studies show person-to-person (P2P) spread can be subclinical or asymptomatic, which might be called “stealth” airborne pathogens because they may infect a substantial portion of the population before resulting in symptoms that cause a visit to a clinic. For example in 2020, of approximately 5000 sailors onboard the aircraft carrier USS Theodore Roosevelt, 55% or 229 of 416 sailors that tested positive for Covid-19 on were asymptomatic [22]. Also in 2021, 18% of high-school classrooms (N=72) in South Africa had Tuberculosis (TB) likely subclinical infection which was discovered only by bioaerosol sampling [23].

Same-day or next-day detection can provide early warning capability needed to break transmission chains of a subclinical, contagious novel bioaerosol cloud in the Pentagon (P2P + NBT). For instance, gain of function (GOF) studies suggest a few gene edits (SNPs) can turn a virus that is ∼50% fatal in dogs into a human virus [24] [25] [26]. If an emerging, airborne measles-relative lacks vaccines or treatments, then a two day delay in its detection results in most of the Pentagon being infected based on the Monte-Carlo simulation of contagion (Figure 3). Since the hypothesized human subvariant has only a small number of SNPs different from the more commonly occuring dog version, expanded sequencing depth in response to detection of a spike is necessary to know the difference.

**Figure 3:**
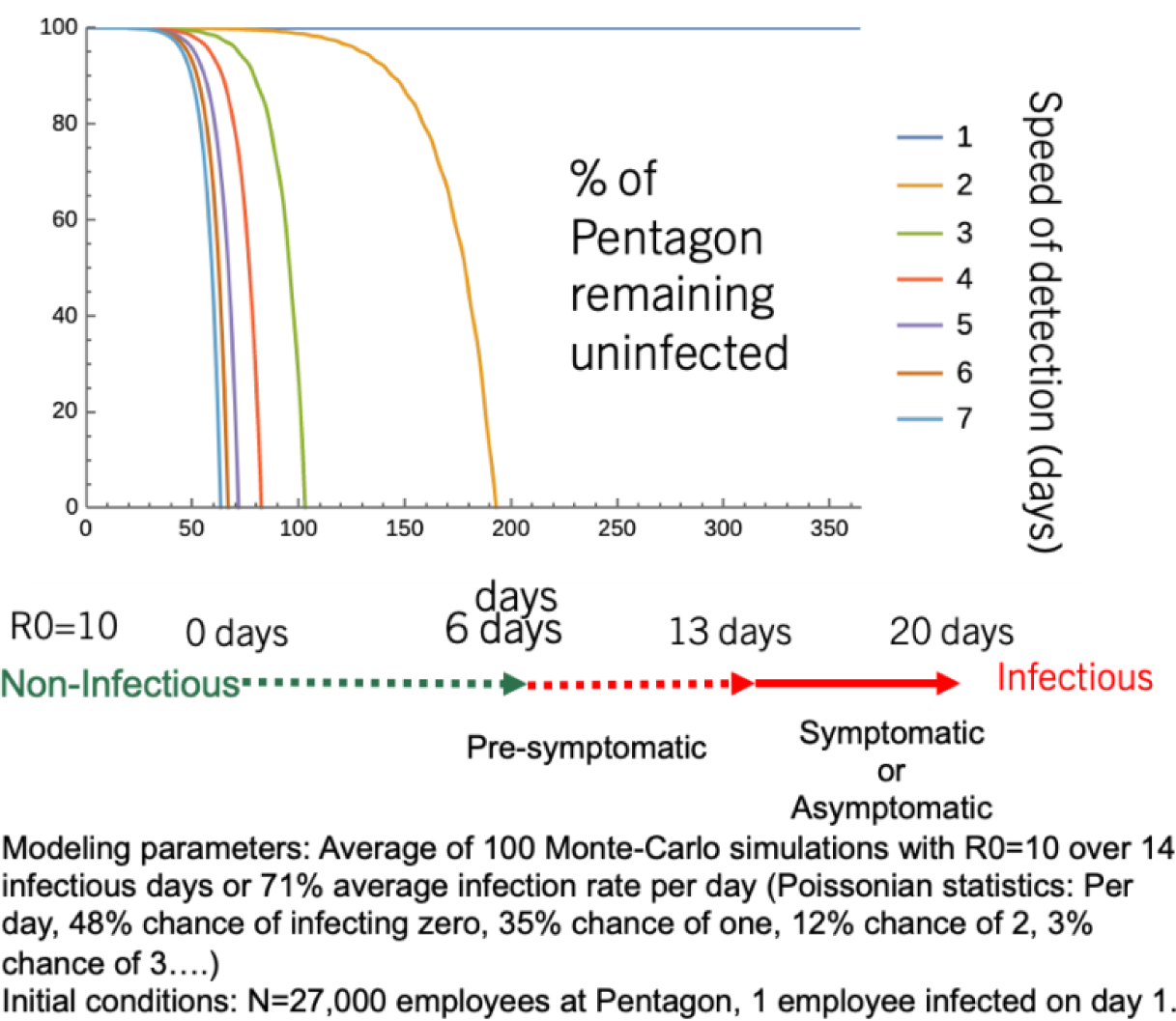
Model of contagion for measles-like virus in the Pentagon.

### Subvariants and Single Nucleotide Polymorphisms (SNPs)

An illustrative model is shown (Figure 4) how increased sequencing depth improves detection accuracy of single-SNP subvariants such as the example canine virus above. It is based on the binomial distribution assuming the fraction of subvariants with that SNP is 1:15 (also known as allelic fraction), and the error rate of the sequencing technology itself that could cause a false read of that SNP is 1:30. A receiver-operator (ROC) curve (Figure 4), true positives versus false positives at different detection thresholds for the SNP, includes different sequencing depths 100, 150, 300, and 1000. In general for true positives to exceed false positives, base error rate must not exceed the allelic fraction of the subvariant being detected. Otherwise the positives would mostly be caused by sequencing errors not actual SNPs.

**Figure 4:**
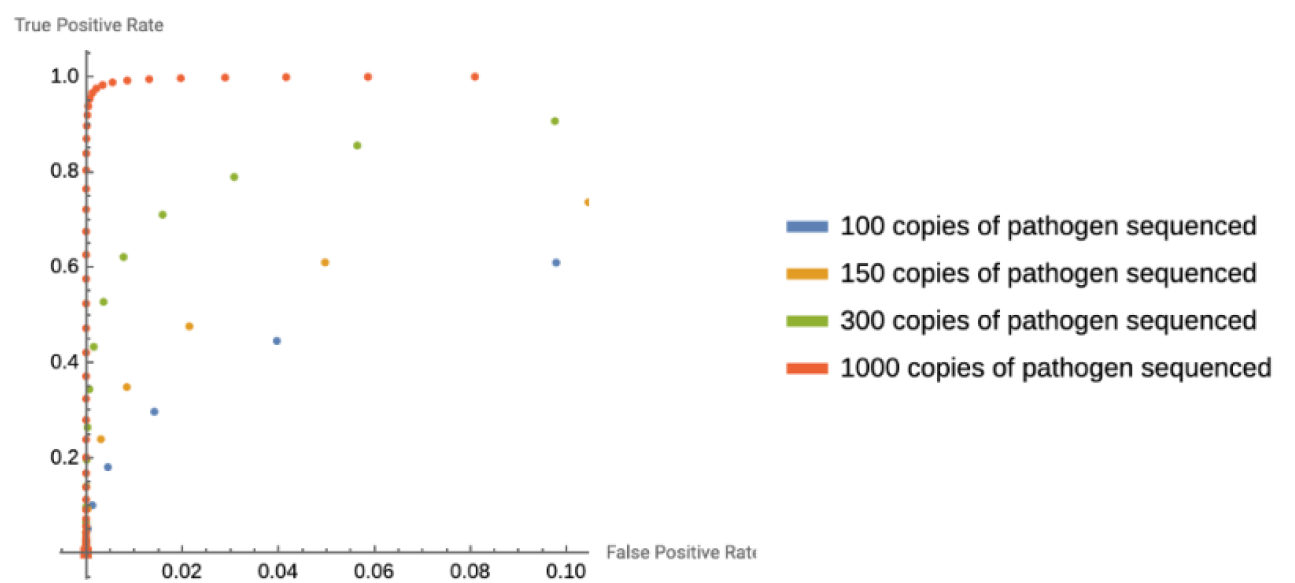
Receiver-operator (ROC) curve for different sequencing depths, 1:30 error rate of sequencing technology and 1:15 ratio of mutated subvariants.

### Detect to Treat Vulnerability (D2T)

Supposing daily metagenomic surveillance were to become feasible for daily operations at the Pentagon (with STV), use of detection still requires sample collection and laboratory processing introducing a delay > 0. This delay necessarily creates a window of vulnerability, between when the incident occurred (“boom”) and when protective actions are taken, right of “boom.” This delay is typically no less than 12-24 hours according to Air Force tactics [10] during which people can still be infected. This residual risk remains with a contagious pathogen (P2P) especially at close contact (Figure 5). The model compares infectious particle concentration in the absence of detection to the reduction seen with 24 hour detection (daily) and immediate use of protection post-detection. This model is based on linear decay in infectious particle concentration with distance from the infectious source derived in [27]. Although assumptions for infectious particles can vary, this model mainly illustrates that daily metagenomic surveillance can substantially reduce risk a day after the initial incident, but cannot alter what transpires between right of “boom” before initial detection of NBT up to or more than 24 hours later.

**Figure 5:**
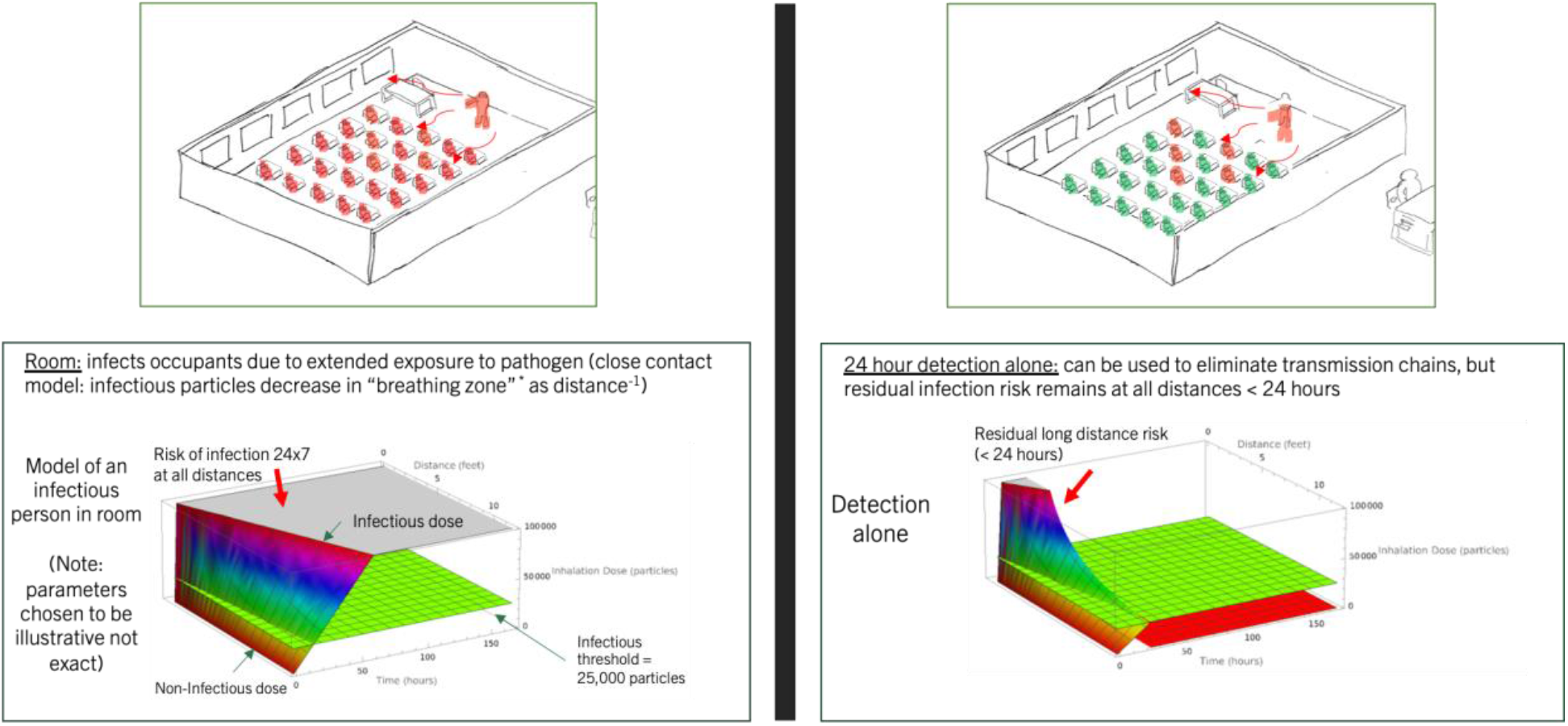
Model comparing infectious particle concentration (P2P) without detection to the reduction seen following 24 hour detection (daily) and immediate use of protection.

To address this window of vulnerability, Air Force tactics describe the primary purpose of DFU network infrastructure deployment, sample collection, and screening of ambient air samples is to provide “Detect to treat” (D2T) notification of a biological attack. D2T is to give time for commanders to recognize the need for, and implement, effective medical treatment protocols. While there are existing treatments for many BSAT, novel treatments for novel threats are unknowable in advance. In 2023, the DoD announced programs using advances in supercomputing-based drug discovery like the discovery of Paxlovid [28], to model new threats and generate potential treatments by modulating the immune system through six metabolic routes [29]. Although Paxlovid for SARS-CoV-2 is a breakthrough, it took years to discover among many alternatives, verify its safety and efficacy in humans, then manufacture, and distribute at first in very limited quantities to the public. It is unclear that such breakthroughs are replicable in time for every emerging bioaerosol threat that comes along in the future. In spite of future biomedical advances, treatments are at high risk of being unavailable for bioaerosol threats for several reasons. For NBT, novel treatments may be unsafe, ineffective, or delayed. For BSAT, treatment-resistant variants of BSAT may emerge [71], or even if they work there simply may not be enough doses or treatment facilities in a surge especially at population-scale (e.g. as seen during the West African Ebola epidemic in 2014 and more recently with COVID-19 and Paxlovid early on after its release). Due to limited manufacturing facilities, treatments are also hard to manufacture on demand during a surge unless stocked in advance.

### Indoor Air Cleaning to “close” the D2T window

Indoors, use of frequent air cleaning 24×7×365 to remove bioaerosols can help “defuse the boom” by cutting the risks during the D2T window of vulnerability since approximately 90% of our time is spent indoors at home, school, work [30]. An 80% lower infection risk (P2P) was observed in Italian classrooms during the Delta/Omicron waves of COVID-19 that were ventilated at the rate of an estimated 5 to 6 air changes per hour (ACH) within the classroom [31] [32]. To use a nuclear analogy, if novel bioaerosols (NBT) are like radioactive air, then its “half-life” (= log_e_(2) ÷ ACH) is inversely proportional to the ACH (Figure 6). The residual level of particle concentration remaining (Figure 6) depends directly on the rate of particle entry or emission into the room, and is also inversely proportional to the ACH [33].

**Figure 6:**
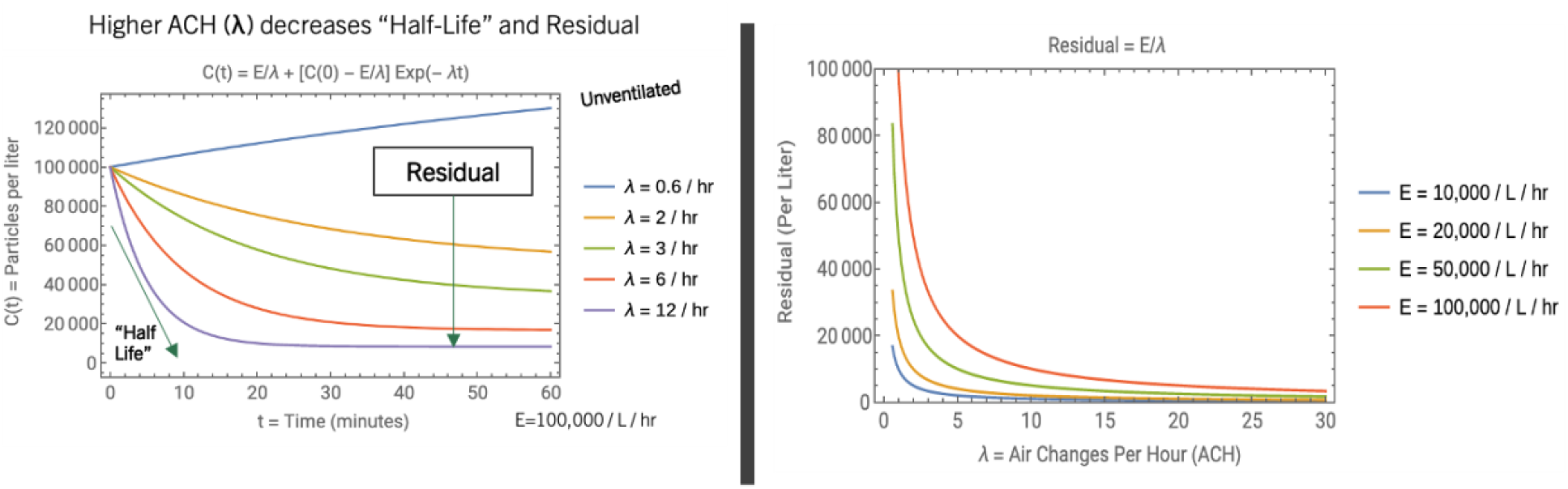
Relationship of Air Changes Per Hour (ACH) to particle concentration.

### 12 Air Changes Per Hour (ACH)

Three data points suggest at least 12 ACH is needed for infection control. First, CDC experts indicated more than 12 ACH (half life < 3.5 mins) is necessary to stop spread of SARS-CoV-2 in some situations although they were not aware of outbreaks in spaces ventilated at 5 to 6 ACH [34] [35]. The volumetric rate of clean air (CADR) needed in a room to meet 6 ACH is 100 cubic feet per minute (100 cfm = 60 × 100 cf per hour) for every 1000 cubic feet (1000 cf). 12 ACH therefore needs double the CADR equal to 200 cfm per 1000 cf.

Second, real-world evidence of the relation of infection risk with high-speed air filtration can be seen with infection (superspreading) rates aboard passenger aircraft. According to CDC, jet aircraft built after the late 1980s recirculate cabin air through filters, and through high-efficiency particulate air (HEPA) filters in most newer-model airplanes [36]. As described below, measurements of the ACH on 3 passenger jets inflight (Airbus A319, A321 and a Boeing 737-Max8/9) were in the range of 11-12 ACH much lower than the 20-30 ACH as reported on the CDC webpage [36], suggesting that perhaps 12 ACH can substantially reduce infection risk from aerosolized pathogens.

Multiple authors observed that superspreading aboard airplanes with high passenger density was less frequent than might be expected with so many passengers contained in such a small space. Most flights are not contact traced, but Australia and New Zealand tracked infections in arriving international flights in early days of pandemic (repatriation) and required passengers to be tested during mandatory quarantine. As of October 20, 2020, out of 62,698 arriving passengers, New Zealand identified 215 who tested positive but published only one instance of inflight super spreading [37]. Australia published three flights with superspreading [38] [39]. If we expect approximately 10% of infected people to be super spreaders [40] about 20 out of 215 arriving in New Zealand and dozens in Australia, well above the published rate of superspreading events. Several researchers have hypothesized why we don ‘t see more superspreading aboard these flights. Some initially credited the institution of mandatory masking for dramatically reduced super spreading aboard flights [41]. However, even prior to mandatory masking aboard aircraft there were relatively few published documented superspreading (mass-transmission) events [42]. Another possible explanation maybe low level of passenger mixing involving close-contact because passengers are confined to seats most of time, yet studies aboard ten transcontinental flights showed there are ample mixing opportunities [43]. If masking or mixing does not account for lower than expected superspreading on flights, a possible remaining explanation is air filtration.

Indoor testing of ACH typically involves generating test aerosols (e.g. salt water, smoke, tracers) which may be unsafe or disallowed e.g. in occupied rooms or passenger aircraft. The procedure to verify ACH on the aircraft relied on ambient aerosols and is based on the model for ambient aerosol decay in the Appendix. At each point in time, the aerosol concentrations (count per liter) were measured using a 7-channel handheld optical particle counter (HOPC) called the Temtop Particle Counter PMD 331 (calibrated to meet ISO-2150). Each measurement was taken for 30 seconds at the most penetrating particle size (MPPS) of 0.3 μm. Since the airplane is an uncontrolled environment several transient aerosol fluctuations can also be observed in aerosol particle counts by the HOPC at MPPS (0.3 μm) after take-off for the three airplanes (Figure 7).

- **Airbus A319:** The best fit (shown) was 11.7 ACH with leak of 3800 particles per liter per hour (32% average percent error).
- **Airbus A321:** The best fit (shown) was 11.8 ACH with leak of 6300 particles per liter per hour (18% average percent error).
- **Boeing 737-Max8/9:** The best fit (shown) was 10.9 ACH with leak of 4100 particles per liter per hour (24% average percent error). Third, in general it takes a reduction of aerosol concentrations by 20 times to get the equivalent of 95% filtration as nominally required for N95 respirators which need at least 12 ACH if not more. Miller et. al. [33] describes an approximation (Equation 3) for equilibrium viral particle concentration (C) with one or more infected persons emitting virus at rate (E per hour) in a well-mixed room of volume (V) with ACH (λ per hour): C = E / (λ V). In an unventilated room (the absence of any indoor ventilation or air filtration), the particle removal still includes surface deposition with a contribution to λ estimated in a wide range of 0.3 to 1.5 [33]. In experiments described in the Appendix the ACH from surface deposition in a room was measured to be approximately 0.6. Assuming surface deposition contributes 0.6 to λ, 20 times reduction in aerosol concentrations to achieve N95-equivalent filtration at far-field in a well-mixed environment requires ACH (λ) from air filtration to be 12.

**Figure 7:**
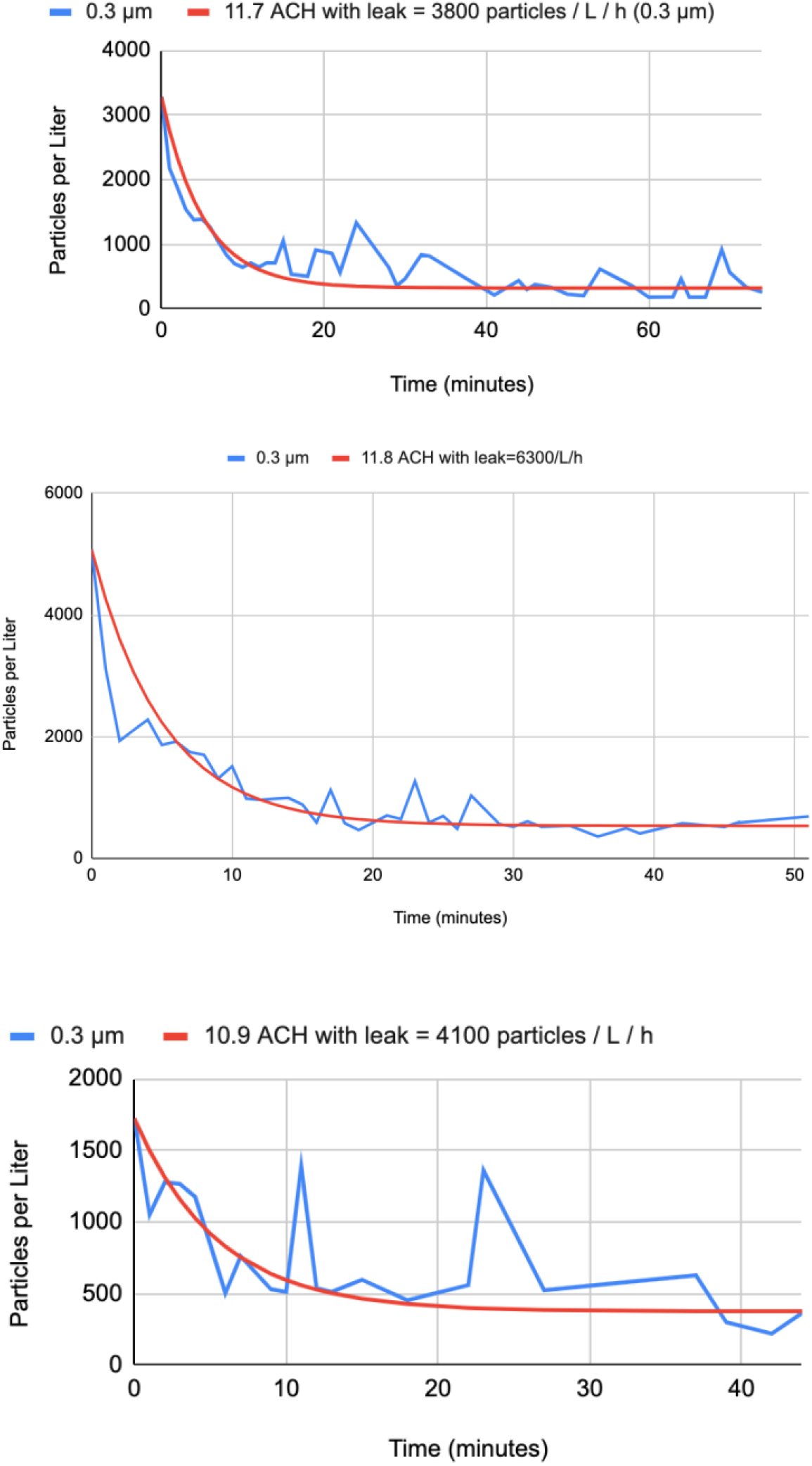
Best fit to estimate ACH after takeoff aboard the Airbus A319, Airbus A321, and Boeing 737-Max8/9.

One caveat is that indoor air cleaning in general is predicted to reduce near-field exposure to exhaled aerosols to some extent [44] dependent on aerodynamic factors [45]. However, a CDC laboratory experiment confirmed [46] that in-room air filtration does not reduce exhaled breath exposure to simulated participants sitting very close to an infected person (near-field e.g. less than 6 feet) to the same degree it can at a larger distance when air is well-mixed within the room (far-field). Respirators remain useful for reducing risk to both near-field and far-field exposures, whereas air filtration can only mitigate far-field exposure in a well-mixed environment, not near-field.

Retrospective analysis showed that use of KN95/N95 respirators by both community members [47] and in healthcare settings [48] resulted in lower COVID infection rates as compared to cloth and surgical masks. Across a variety of manufacturers the average filtration efficiency of N95 at\ 0.3 μm particle size was found to be much higher at 98% and more consistent than 81% for KN95 (Table 2 of [49]). This 98% is reassuringly higher than the 95% filtration efficiency nominally required for N95, although N95 fit [50] and filtration efficiency (Figure 3 of [51]) may degrade with reuse over several hours (and wearing an N95 with poor fit is like closing the windows of a car door but leaving a crack open). The N95 average of 98% requires a 50 times reduction in inhaled aerosols (ACH=30) and KN95 average of 80% requires 5 times (ACH=3) in an unventilated room assuming surface deposition contributes 0.6 to ACH (λ).

### Portable Air Filtration

It is not possible to protect from NBT or BSAT using outside air for indoor ventilation if the source of the bioaerosol release is from outside, unless the outside air is nearly 100% disinfected or filtered when entering the building, which is often cost-prohibitive. Infection rate of a simulated anthrax release from any localized source (e.g. point or line) has distance-dependent dilution of bioaerosols as modeled in Figure 3 (b) of [78]. Below a certain threshold, infection risk decreases sharply when inhaled dose is reduced by an order of magnitude or more as shown in Figure 3 (a) of [78].

With some exceptions, more than 4 ACH of filtered, recirculated air using centralized heating ventilation and air conditioning (HVAC) systems is also uneconomical because of the cost of energy needed to transport the air remotely to an HVAC rises approximately as the cube of ACH (ACH^3^) [52]. This is not including the costs of conditioning the air temperature at these higher airflow rates.

To push above 4 ACH to 12 ACH and beyond, in-room air cleaning methods such as air filtration and germicidal ultraviolet (GUV) technology can be used economically. The costs of recycling the indoor air by locally cleaning air scales up linearly proportional to target ACH (ACH^1^) and the room ‘s volume. Public Health authorities including CDC [53], EPA [54], White House [55] [56], and California Department of Public Health (CDPH) [57] recommend installation of portable air filtration, HEPA or Do-It-Yourself (DIY), for use in all schools, homes, businesses. HEPA air purifiers are widely available in retail channels and in widespread use in US households to cut down ambient aerosol concentrations not only from infectious pathogens such as SARS-CoV-2, Influenza, RSV but also allergens, cooking pollution (frying, toasting, etc), wildfire particulates, wood-burning particulates, road pollution (diesel soot), and industrial pollution.

In one instance (Figure 8) ambient particle counts at 0.3 μm (most penetrating particle size) in a room within a California home were reduced by almost 50x from outdoors to indoors within 40 minutes after closing the windows and running 9 ACH of DIY air filtration. This is easy to replicate except near bioaerosol sources or close contact with infectious people (Figure 9).

**Figure 8:**
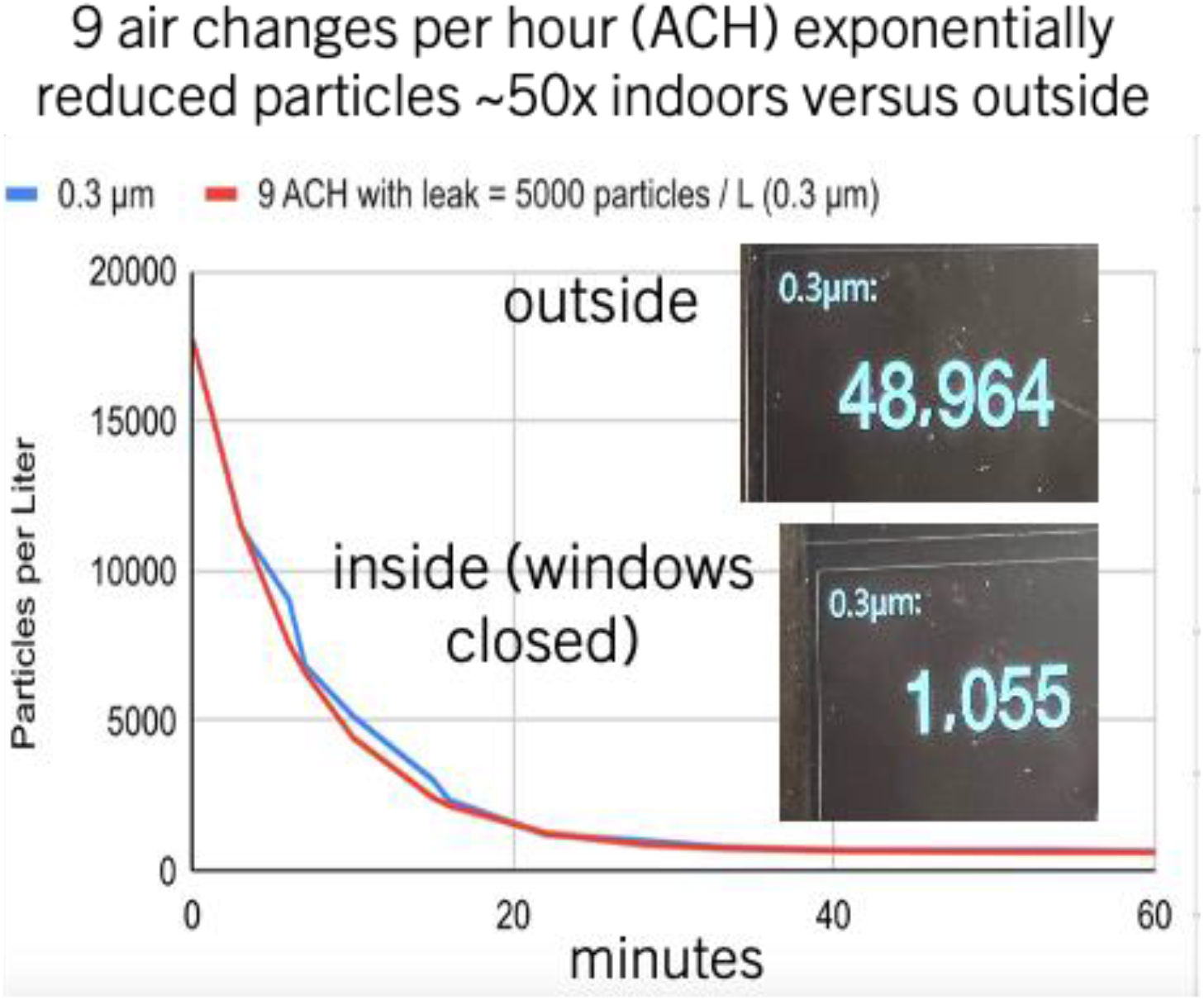
With 9 ACH air filtration indoors, particle concentrations were reduced by almost 50x relative to outside.

**Figure 9:**
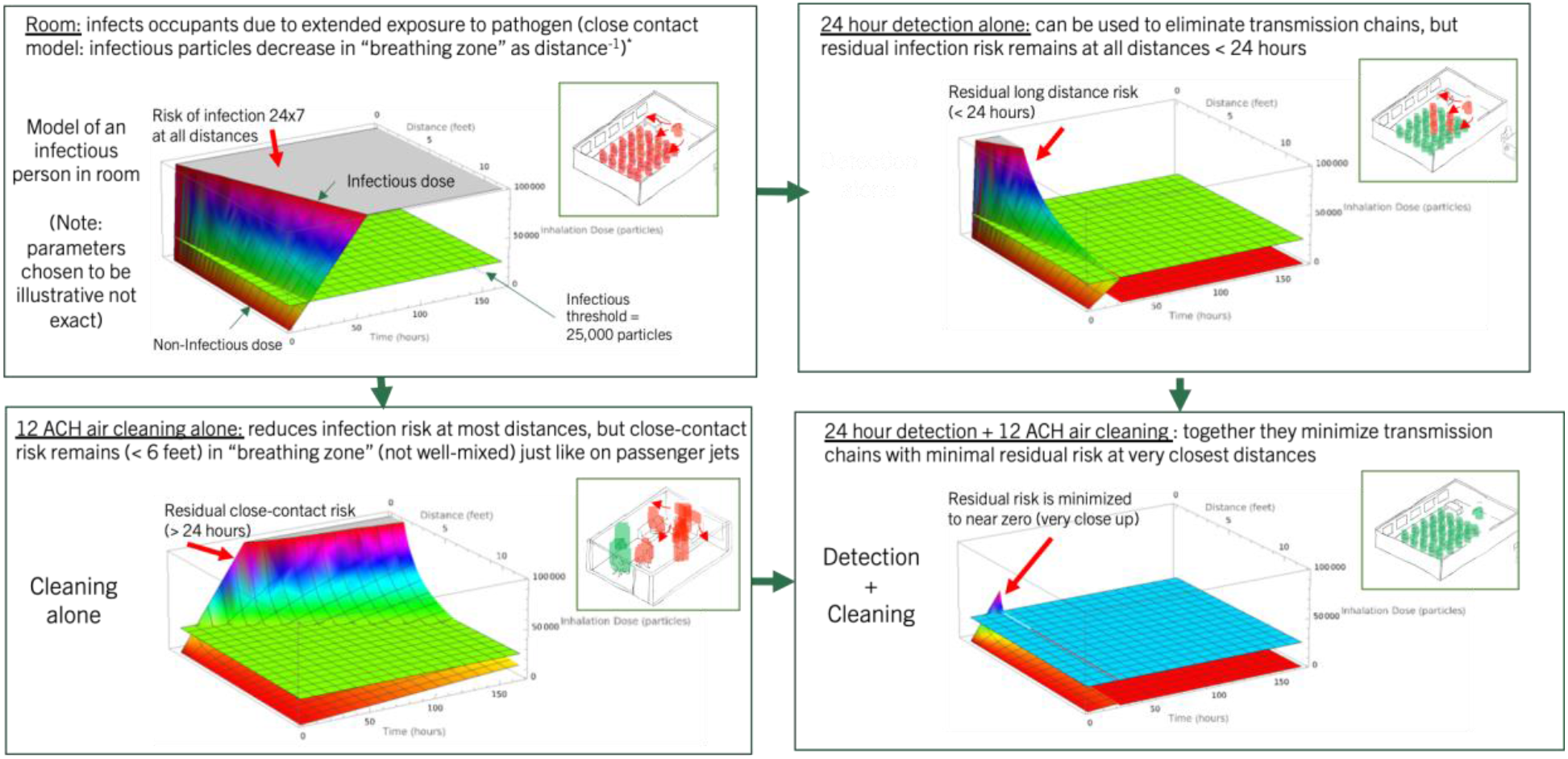
Infectious particle concentration (P2P) without detection to reduction following 24 hour detection (daily), 12 ACH air filtration (24×7×365), and both in combination.

GUV has been in use in many applications across DoD [73], in hospitals for infection control and to some extent for short-term or temporary exposure in public spaces. At this time a debate remains unresolved about long-term safety and appropriateness for extended, daily use of GUV [58] especially in regards to excessive generation of volatile organic compounds (VOC) and formation of secondary aerosols [59] [60] which may be carcinogenic [61].

### Combination of Detection and Air Cleaning

With continuous indoor air cleaning to reduce bioaerosol concentration, treatment is not the only option upon detection of a bioaerosol incident (NBT). Post-detection, exposure to bioaerosols can be further reduced by establishing emergency protocols for use for the duration of the detected disaster-incident. These temporary measures include well-fitting, comfortable respirators [62] not just cloth masks (e.g. N95, N99, P100, etc.) both outside and inside, and optionally with temporary indoor use of GUV and higher speed air filtration. The model of Figure 5 is used to estimate the combination effect on reduction of infectious particle concentration (P2P contagion) using both 24×7×365 detection with 12 ACH air filtration in the room (Figure 9). In the model, detection alone allows a window of infection (delay > 0) and filtration (or UV) alone still allows infection in the “breathing zone,” but in combo they minimize risk (from P2P + D2T). At each distance from the bioaerosol source (infected person), the air filtration reduces exposure to the bioaerosol until detection of the incident (during the D2T window), and subsequently post-detection risk is minimized due to additional protections or treatment that maybe used.

### High-Efficiency Particulate Absorbing (HEPA) and Do-It-Yourself (DIY) Air Cleaners

Do-It-Yourself (DIY) air filtration uses HVAC filters rated MERV 13 or higher in conjunction with box fans to achieve indoor air filtration results comparable to HEPA filtration but up to 10x lower cost [63]. To reach > 10 ACH in rooms with typical 8 ‘-10 ‘ ceiling heights, HEPA purifiers running at tolerable noise levels cost in the range of $2-$5 per square foot whereas Do-It-Yourself (DIY) air purifiers cost $0.5 to $1 per square foot, verified in tests described below. In 2022, CDC (NIOSH) conducted a “crash test” of DIY air purifiers using aerosols at the most penetrating particle size (0.3 μm to 0.6 μm), to verify whether they are capable of stopping viruses between test dummies [64]. In these tests, CDC found 12+ air changes per hour (12 ACH) was achievable in a realistic size classroom of 6000+ cubic feet but it took more than one DIY air purifier (parallelization). In 2021, EPA-sponsored testing of DIY by Underwriter Labs found popular box fan models remained safe after stress tests with loaded and blocked filters [65]. DIY air purifiers not only reduce bioaerosol risk, improve air quality in general, but they may also double as low-cost bioaerosol collectors in lieu of DFUs [66].

Using similar methods as measurements of ACH in the passenger airplanes above, the ACH derived CADR is estimated below (Table 3) for a variety of HEPA and DIY models tested using a uniform methodology relying on ambient aerosols. DIY models were assembled using the instructions provided in [67]. As a general trend, the cost of 1000 cfm is significantly lower for the DIY configurations ($279-$699) than the HEPA ($334-$1300). Noise is an important aspect for end-user acceptance, and 20” box fans made by Lasko running at their lowest speed (as tested below) are widely used in classroom applications. The HEPA were tested at their highest speed. Notably the lowest noise for DIY used ten 14 cm PC fans (“Tower of Power”, 429 cfm). The lowest noise HEPA was the SmartHealth Blast, 829 cfm. Both had comparatively high levels of CADR compared to most other models. A typical classroom of size 10,000 cubic feet would require 1000 cfm to reach 6 ACH, and 2000 cfm for 12 ACH. Of these models, the lowest cost approach would be to use the DIY box fan (Lasko) with 2” MERV 13 from Nordic Pure as assembled in [67] and tested in [63] which for typical 10 ‘ ceiling heights translates to approximately $0.5 per square foot to get > 10 ACH. The cost per square foot would rise proportionally for other models based on the cost per 1000 cfm below.

**Table 3:** Performance characteristics of DIY and HEPA air purifiers.

| Purifier | Type | ACH-Derived<br>CADR (cfm) @<br>0.3 $\mu\text{m}$ | Cost | CADR<br>(cfm) per<br>\$100 | Cost (\$) of 1000<br>cfm | Noise<br>at 9"<br>(dBA) | Weight<br>(lbs) | Volum<br>e<br>(cubic<br>feet) |
| --- | --- | --- | --- | --- | --- | --- | --- | --- |
| Nordic Pure 2" MERV 13 (w/ box fan) @ speed low | DIY | 197.4 | \$55 | 358.91 | 278.6220871 | 58 | 8.00 | 1.8 |
| Nordic Pure 2" MERV 13 4-filter cube (w/ box fan) @ low | DIY | 310.905 | \$100 | 310.91 | 321.641659 | 57 | 8.00 | 6.60 |
| 3M 1" MERV 14 (w/ box fan) @ low | DIY | 212.205 | \$70 | 303.15 | 329.8697015 | 58 | 8.00 | 1.50 |
| Taotronics TT-AP003 @ speed 3 | HEPA | 236 | \$79 | 299 | 334.4481605 | 58 | 13 | 1.4 |
| Lasko Airflex MERV 10 (w/ box fan) @ speed low | DIY | 162.855 | \$69 | 236.02 | 423.6897854 | 55 | 11.00 | 1.70 |
| Lennox 5" MERV 16 (w/ box fan) @ speed low | DIY | 330.645 | \$160 | 206.65 | 483.9026751 | 58 | 10.00 | 2.40 |
| Tower of Power, 3M 1" MERV 13 (w/ 10 Arctic P14 PC fans) | DIY | 429 | \$300 | 143 | 699.30069<br>93 | 50 | 10 | 2.9 |
| Levoit Core 600S @ speed 4 | HEPA | 419.475 | \$299 | 140.29 | 712.79575<br>66 | 66 | 14.00 | 2.00 |
| Honeywell HPA-5100B @ speed Turbo | HEPA | 172.725 | \$129 | 133.90 | 746.85193<br>23 | 62 | 14.00 | 1.10 |
| Coway AP-1216L @ speed 3 | HEPA | 227.01 | \$179 | 126.82 | 788.51916<br>1 | 65 | 13.2 | 1.7 |
| Smart Health Blast @ speed max | HEPA | 829.08 | \$999 | 82.99 | 1204.9500<br>65 | 53 | 69 | 6.4 |
| Coway Airmega 400 @ speed 3 | HEPA | 379.995 | \$494 | 76.92 | 1300.0171<br>05 | 63 | 25.00 | 3.00 |

### Filter Replacement Cycle

With air filtration products the filters will need to be replaced at some point after they have been used. Below are measurements (tests) of the filtration efficiency for 5” Lennox MERV 16 filters used in DIY air purifiers at the MPPS of 0.3 μm. Each dot (Figure 10) represents a different filter instance and the x-axis was ordered in ascending order of filtration efficiency (not time). These filters were used for approximately 6 months and compared to fresh (new) filters installed in a school (Figure 10). The usage pattern was 5 days per week during school hours. The chart shows that some filters appear to have lower filtration efficiency than others possibly due to gradual loss of electrostatic charge since these are electret filters. The lower the efficiency the lower the clean air delivery rate, although the efficiency typically remains higher at larger particle diameters. Depending on the chosen threshold for replacement, policies for replacement intervals can be chosen based on testing the filters individually, or they may be replaced periodically (e.g. 1 year) without testing each one. Generally HEPA filters are expected to retain filtration efficiency longer than HVAC filters but actual results will vary based on a number of usage and environmental conditions.

**Figure 10:**
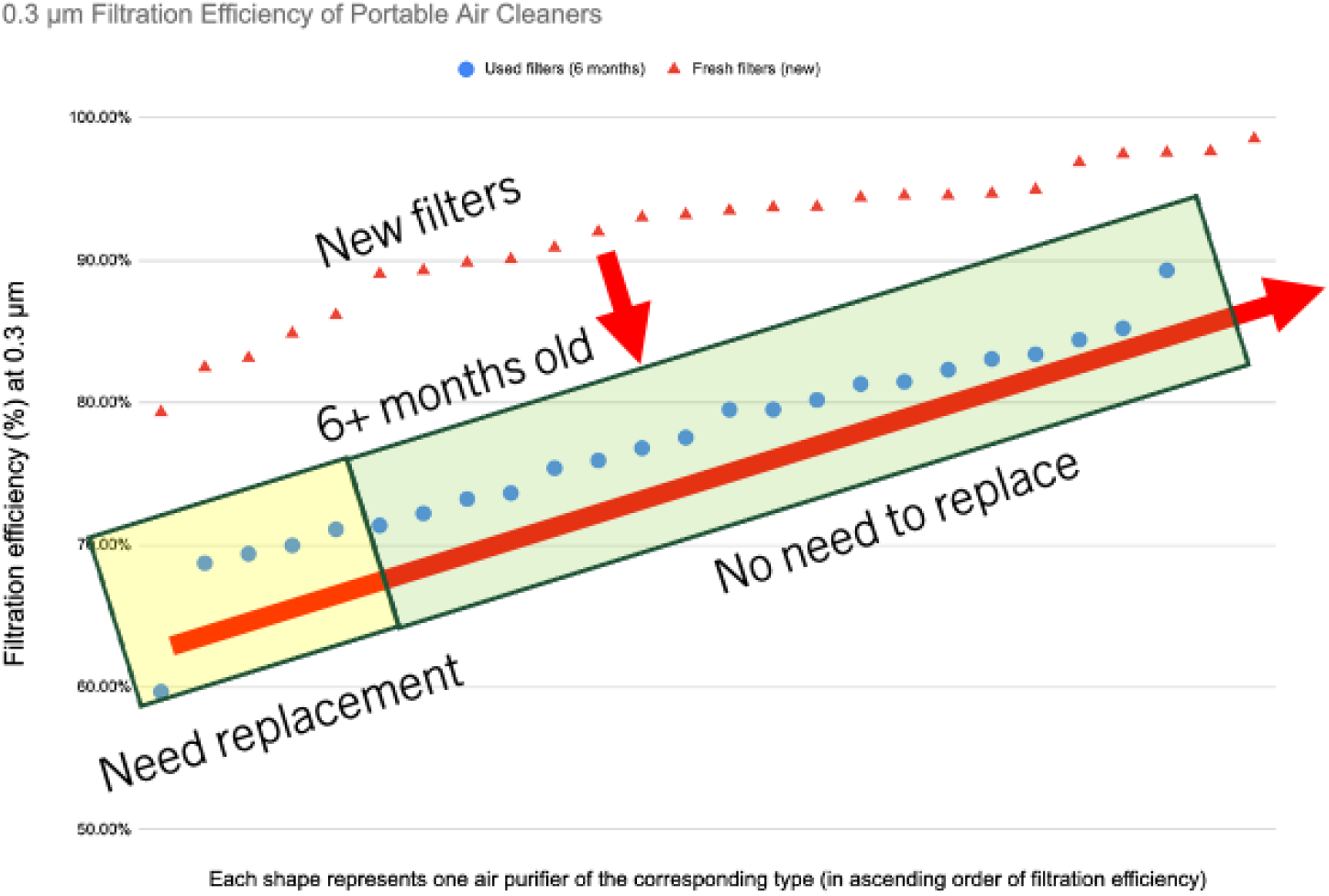
Filtration efficiency of DIY air filters in use for 6 months compared to new filters.

### Conclusion and Next Steps

The PFPA pilot with N=105 DFUs at the Pentagon reservation highlighted economic barriers to adoption of metagenomics for emerging bioaerosol threat detection on a daily or routine basis. If the Pentagon is experiencing these economic barriers, they are likely to be experienced at many other secure DoD facilities as well as civilian buildings around the world. These barriers included the reagents used for metagenomics being two orders of magnitude more costly than PCR, in addition to the cost of sequencing instruments (machines) and the labor (expertise) to analyze the bioinformatics. These economic barriers can be addressed in principle by dynamically pooling of samples from the network of DFUs called Spike Triggered Virtualization (STV), whereby the pooling factor and sequencing depth are modulated automatically based on novel biothreats in the sequencing output. By running at a high enough average pooling factor, the daily and annual cost per DFU can be reduced by one to two orders of magnitude depending on the trigger thresholds chosen.

It is unlikely, a covert aerosol release of a biological agent would only be captured by a single DFU, but the possibility exists. The risk of this possibility can be mitigated with deployment of more DFUs in the network. In case of contagious pathogens exhaled by infected people such as SARS-CoV-2, “spikes” of bioaerosol can be observed not only as a result of a rising number of infected people (temporally correlated “waves”) but are likely because bioaerosol emissions can vary by one or two orders of magnitude due to physiological differences among infected people (spatially correlated). For contagious pathogens, a Monte-Carlo model of contagion in the Pentagon suggests same-day or next-day detection is necessary to provide early warning capability to break transmission chains. The sensitivity of the trigger for detection of pathogenic novelty in STV depends on the chosen threshold concentration to detect the spike. There are opportunities to enhance the sensitivity of the STV trigger further with artificial intelligence (AI).

Provided detection within 24 hours is achievable, the “Detect to Treat” doctrine for BSAT (known threats) becomes a risk with novel bioaerosol threats because novel treatments either may not exist or if they do they may still be hard to come by. Exposure to bioaerosols post-detection can be reduced by establishing protocols, for the duration of the detected incident, to use well-fitting, comfortable respirators not just cloth masks (e.g. N95, N99, P100, etc.) both outside and inside, optionally with temporary indoor use of GUV and higher speed air filtration.

People can still be infected during the 24 hour window between a bioaerosol incident and its detection (D2T gap). This infection risk during the D2T gap can be reduced economically with high-speed indoor air cleaning of more than 10 air changes per hour (10 ACH) similar to the ACH level observed on passenger airplanes inflight. 4 ACH or more is likely to be cost-prohibitive using central HVAC systems, but 10 ACH or more can be achieved economically using portable air filtration in rooms with typical ceiling heights (< 10 ‘) for a per square foot cost of approximately $0.5 to $1 for Do-It-Yourself (DIY) and $2 to $5 for HEPA.

Next steps. Building on the groundwork in the Pentagon ‘s pilot study, a DFU network infrastructure in a campus or office at the similar scale as the Pentagon ‘s pilot (N=105) is needed to verify economical daily operations of using metagenomics for detection of novel bioaerosol threats, based on STV and AI-enhanced triggers to detect spatio-temporally correlated spikes. One test case for the detection system would be successfully tracking the rise and fall of SARS-Cov-2 spikes (waves) using STV, efficiently modulating sequencing rate of SARS-CoV-2 variants and their location within the campus. Simultaneously, 12 ACH indoor air filtration throughout the campus using HEPA or DIY closes the gap between biothreat release and its detection in DFUs to secure from emerging bioaerosol threats. The viability of air purifiers used for high-speed air filtration (as complementary to DFUs) to sample the air for aerosolized pathogens needs to be tested and verified.

## Data Availability

All data produced in the present study are available upon reasonable request to the authors

## Funding

No external funding was received.

### Appendix

Ambient aerosol decay with air filtration in enclosed indoor space

The mathematical model for aerosol concentration from an infected person in a room that was described in [33] can be adapted to measure ACH in a whole house or room after any ambient (or generated) aerosol has been introduced. The differential equation (Equation 2 in [33]) is the same, but in contrast to the rising solution that asymptotically approaches C = E/λV given in [33], the ambient aerosol concentration in the room or house at time t after windows are closed can be expressed as the alternate (decaying) solution to the same differential equation:

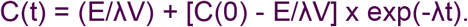

where

C(t) = ambient aerosol concentration at time t

C(0) = initial concentration

E = rate of ambient aerosols “leaking” into room or house (per hour) e.g. from outside

V = volume of room or house

λ = ACH

exp = exponential function.

### Procedure

To measure the ACH of air filtration (HEPA or DIY) in an indoor space such as a room, first ambient aerosols from outdoors are allowed to enter the indoor space by opening windows and doors and get close to the level outdoors as measured by the optical particle counter described below. Then the windows are closed, and then finally the air filtration is turned on to track the decay of indoor aerosol concentrations with an optical particle counter. The shape of the decay curve and asymptote reveal the ACH and leaks of outdoor aerosols into the room or whole house. In case of an airplane, such control is not possible in which case the ambient aerosol concentration is determined by the opening/closing of doors by the crew, the entry of outdoor aerosols through airframe leaks or air intake in the cabin, and other in-cabin aerosol generating events.

### Measurement of noise generated by air purifiers

Noise was measured for each air purifier/filter using an iPhone app [68] maintained by The National Institute for Occupational Safety and Health (NIOSH) at a 9″ distance perpendicular to the direction of the output airflow.

### Estimation of ACH and rate of indoor leakage

All measurements (counts) were input into Google Sheets and compared to the model for ambient aerosol decay with different values of ACH and indoor aerosol leak rate. The aerosol concentration measurements representing C(t) are fitted by a computer program (written in Python) to obtain the best approximation for ACH (λ) and the rate of indoor leak (E/V) at 0.3 μm particle size by minimizing sum of percent error across each measurement (in increments of 0.1 ACH and 100 particles per liter per hour). For example, (Figure 11) the best fit (2.7% average percent error) for surface deposition in the room with windows closed and without any filtration or HVAC enabled was with 0.6 ACH and leak equal to 4900 particles per liter per hour. The plot (Figure 11) of the best fit (4.3% average percent error) obtained for the Coway Airmega 400 in the Room with 7.7 ACH and leak equal to 4700 particles per liter per hour.

**Figure 11:**
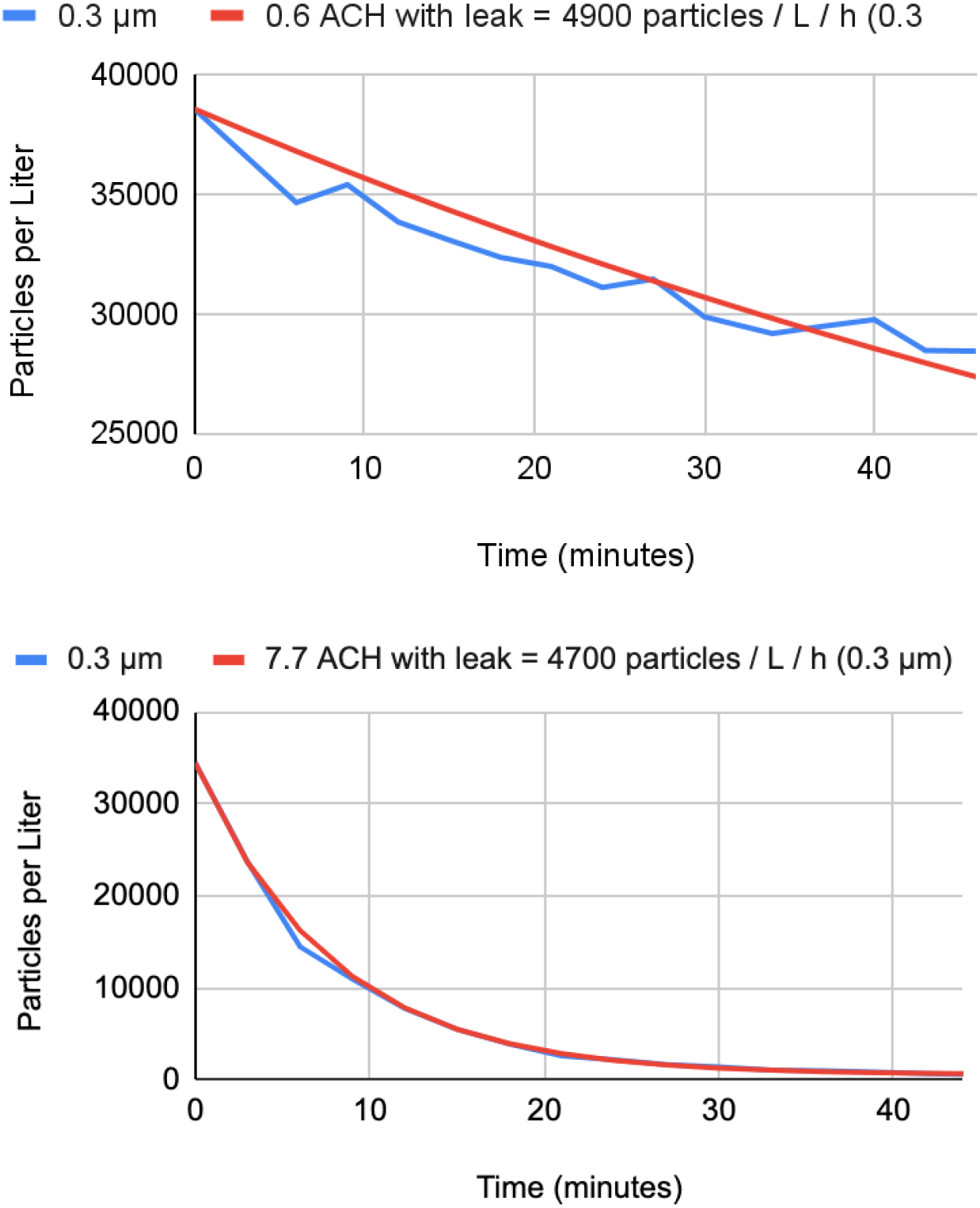
Example of best fit with 0.6 ACH the rate of surface deposition in Room, and best fit with 7.7 ACH for Coway Airmega 400 in Room.

Similarly (Figure 11), the ACH estimates for each of the purifiers tested and their average percent error is listed (Table 4). Since the measurements were taken at different times and ambient aerosols outside vary from day to day, the leak was also observed to vary between tests. The tests were conducted in a representative room of estimated size 2961 cubic feet. Tests using particle decay rates were conducted by EPA for a variety of DIY air purifier configurations [69].

**Table 4:** ACH estimates for each of the HEPA / DIT air purifiers tested.

| **Purifier** | **Type** | **ACH (per hour)** | **Average Percent Error (18 data points)** | **Leak (particles / L / hour)** |
| --- | --- | --- | --- | --- |
| Surface (Room) | n/a | 0.6 | 2.70% | 4900 |
| Lasko Airflex MERV 10 (w/ box fan) @ speed low | DIY | 3.3 | 2.10% | 21700 |
| Honeywell HPA-5100B @ speed Turbo | HEPA | 3.5 | 3.70% | 12900 |
| Nordic Pure 2" MERV 13 (w/ box fan) @ speed low | DIY | 4 | 2.70% | 6500 |
| 3M 1" MERV 14 (w/ box fan) @ low | DIY | 4.3 | 2.90% | 11000 |
| Coway AP-1216L (speed 3) | HEPA | 4.6 | 5.70% | 300 |
| Taotronics TT-AP003 @ speed 3 | HEPA | 4.8 | 6.17% | 30300 |
| Nordic Pure 2" MERV 13 4-filter cube (w/ box fan) @ low | DIY | 6.3 | 3.70% | 9800 |
| Lennox 5" MERV 16 (w/ box fan) @ speed low | DIY | 6.7 | 2.80% | 11500 |
| Coway Airmega 400 @ speed 3 | HEPA | 7.7 | 4.30% | 4700 |
| Levoit Core 600S @ speed 4 | HEPA | 8.5 | 6.90% | 2300 |
| Tower of Power, 3M 1" MERV 13 (w/ 10 Arctic P14 PC fans) | DIY | 8.7 | 8.70% | 1900 |
| Smart Health Blast @ speed max | HEPA | 16.8 | 9.40% | 10100 |

